# Polygenic score for sleep duration in relation to risk of Alzheimer’s disease: results from the UK Biobank

**DOI:** 10.1101/2022.12.15.22283413

**Authors:** Angel TY Wong, Sarah Floud, Gillian K Reeves, Michael V Holmes, Ruth Travis, Cornelia M van Duijn, Aiden Doherty, Karl Smith-Byrne

**Affiliations:** Cancer Epidemiology Unit, Nuffield Department of Population Health, University of Oxford; Medical Research Council, Integrative Epidemiology Unit, University of Bristol, Bristol; Nuffield Department of Population Health, University of Oxford

**Author notes:** Corresponding author: Dr. Aiden Doherty. joint last authors (Aiden Doherty, Karl Smith-Byrne).

**Keywords:** Alzheimer’s disease, sleep duration, polygenic score, dementia

## Abstract

**INTRODUCTION:** Studies have suggested sleep duration may be associated with Alzheimer’s disease risk, but findings based on self-reported sleep duration are likely to be influenced by reverse causation and residual confounding bias.

**METHODS:** A polygenic score (PGS) for device-measured sleep duration was constructed using LDpred2-auto in 77,770 white British UK Biobank participants. We applied the PGS to 264,746 white British participants independent of the sample from which the PGS was developed. We assessed the association of fifths of genetically predicted sleep duration with Alzheimer’s disease risk (1,451 cases/264,746 individuals over median 12.5y of follow-up).

**RESULTS:** The PGS explained ∼2% of variation in device-measured sleep duration. Compared to individuals in the middle fifth of PGS, those in the highest fifth (indicating ∼15 mins/day longer sleep) had a lower risk of Alzheimer’s disease (HR=0.79[95%CI,0.67-0.94]).

**DISCUSSION:** Our results indicate that genetic predisposition to relatively long sleep duration is associated with a lower Alzheimer’s disease risk.

## Introduction

Poor sleep is a proposed risk factor for dementia [1] due to, for example, increased beta-amyloid burden after acute sleep deprivation [2]. Existing studies have typically reported the association of self-reported sleep duration, which may be subject to some degree of measurement error and be influenced by early symptoms of Alzheimer’s disease thus leading to reverse causation. Moreover, a meta-analysis reported evidence for long sleep as a potential risk factor for dementia [3]. With over half of the included prospective studies having fewer than 10 years of follow-up [3], it is difficult to exclude that the association is a consequence of dementia pathology in a prodromal stage (reverse causation).

Sleep duration from accelerometers may provide a more reliable measurement of sleep by reducing measurement error. Further, polygenic scores (PGS) based on genome-wide association studies (GWAS) can provide highly informative predictors of complex traits, such as sleep duration, that are less likely to be affected by reverse causality. Thus far, GWAS have identified few single-nucleotide polymorphisms (SNPs) for device-derived sleep duration [4,5], which explain limited trait variance (0.33%)[4]. Given GWAS sample sizes (n∼91,000) and the limited variation explained by previously identified significant SNPs, investigation of sleep duration and dementia risk using Mendelian randomisation (MR) may be underpowered, as reflected by the wide confidence intervals around the estimates in previous studies [6-8]. We have previously reported that up to 18% of the phenotypic variance could be explained by common SNPs (P<5-e-3) [4], so we hypothesise that a PGS based on this wider range of genetic variants may increase power to better investigate the role of sleep duration in dementia risk.

This study aimed to assess the association between a new PGS for device-measured sleep duration and Alzheimer’s disease risk in UK Biobank participants.

## Methods

### Study population

UK Biobank recruited ∼500,000 individuals living in UK aged 40-69 years in 2006-2010 [9]. Ethical approval for the study was granted by The National Information Governance Board for Health and Social Care and the North West Multicentre Research Ethics Committee [9]. All participants provided electronically signed consent. At recruitment, participants answered a touch-screen questionnaire, provided biological samples, underwent an anthropometric assessment, and were interviewed by a trained nurse. We used the full genotype data release [10] (∼96 million SNPs) for ∼480,000 individuals (genotyped and imputed variants based on the UK10K haplotype, the 1000 Genomes Phase 3, and the Haplotype Reference Consortium reference panels) [4].

### Study design

The study design is shown in Figure 1. We first conducted a GWAS for device-measured sleep duration among UK Biobank participants with accelerometer data, then re-estimated the effect sizes using the LDpred2-auto algorithm, and applied the resulting PGS for sleep duration to an unrelated sample in the cohort in order to assess the association of genetically predicted sleep duration with Alzheimer’s disease.

**Figure 1.**
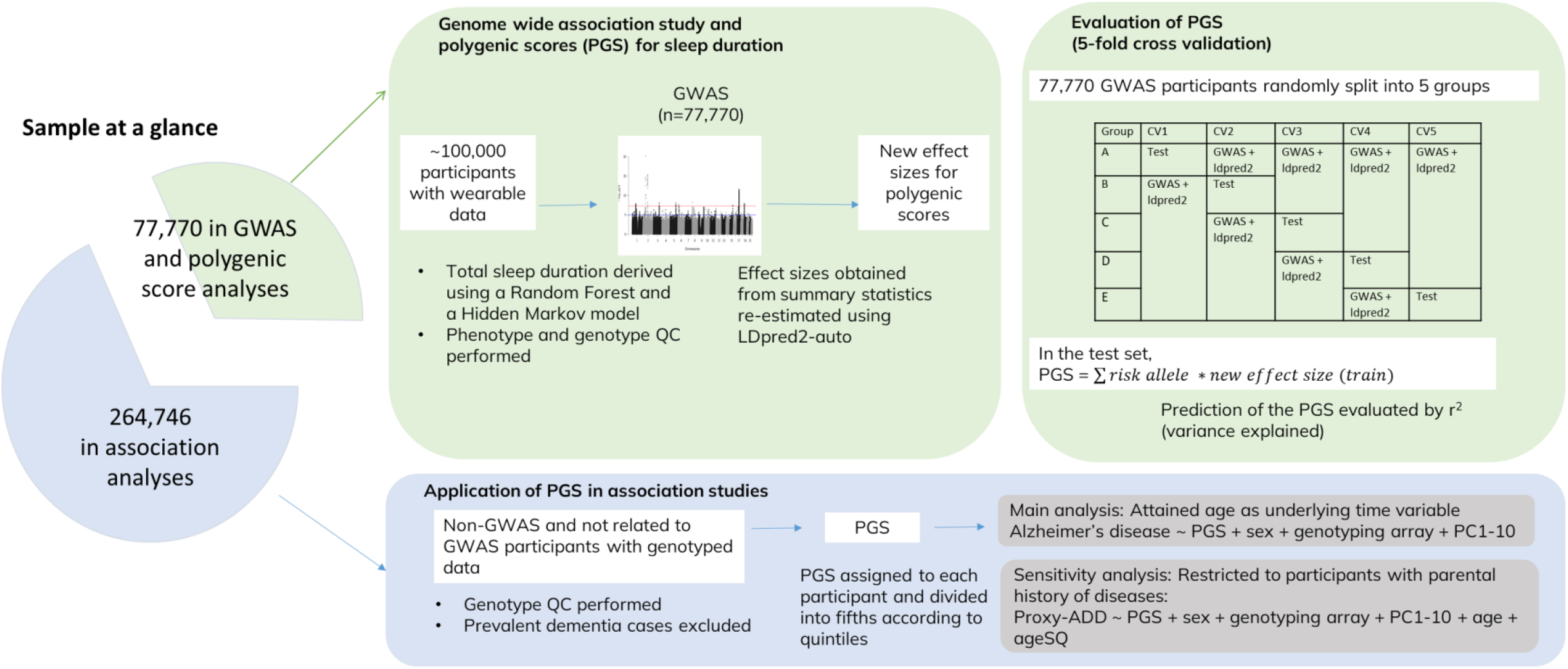
Study flowchart to investigate the association between the polygenic score for sleep duration and Alzheimer’s disease risk

### Polygenic risk score for sleep duration

#### Measurement of sleep duration: Accelerometers

After recruitment, ∼100,000 participants were asked to participate in a wrist-worn accelerometer data collection in 2013-2015. They wore an Axivity AX3 on their dominant hand at all times for a 7-day period [11]. We measured sleep duration from the device using the previously published random forest and hidden Markov model-based approach [11], with details described elsewhere [4,11,12]. In this analysis, average sleep duration during the period that the device was worn (hereafter called sleep duration) was the exposure, and the unit was converted from ‘proportion over time’ to ‘hours of sleep per 24 hours’.

#### Genome-wide association analysis for sleep duration

Among 103,640 adults who wore activity monitors, we excluded 7,044 participants with missing or poor wear time (<3 days wear or did not have wear in each hour of the 24 hour day), 3 participants with >1% of values clipped before or after calibration (i.e., falling outside the dynamic range +/-8g of the accelerometer [11]), and 14 participants with no accelerometer value or unrealistically high average accelerometer values (average vector magnitude >=100 mg).

Of the 96,579 remaining participants, 94,385 had genotype data. We removed 55 participants who had different self-reported and genetically inferred sex and 57 participants who had putative sex chromosome aneuploidy. The phenotype and genotype quality control procedures broadly followed Doherty et al. (2018) [4]. As we restricted our sample to genetically white British participants and an unrelated sample, we excluded 13,057 participants who were not of white British genetic ancestry, 34 participants who had excess relatives, and 3,412 participants who had at least one 3^rd^-degree relative in the remaining sample using the relatedness file [10], leaving 77,770 participants in the GWAS. We only analysed autosomal SNPs, and excluded SNPs with MAF <1% and imputation score R^2^ <0.3.

We performed linear mixed model analysis under an infinitesimal model implemented in BOLT-LMM program (version 2.3) [13] for the GWAS of sleep duration, adjusting for genotyping array, season of wear (spring, summer, autumn, winter), assessment centre, genetic sex, and the first 10 principal components of ancestry, age at recruitment, and age-squared. We adjusted for season of wear because a previous analysis reported differences in sleep duration in summer versus winter in the enhancement cohort [12]. We used Plink (version 2) [14] to exclude variants that deviated from Hardy-Weinberg equilibrium (p<1e-7). The Manhattan plot and QQ plot were plotted using qqman [15]. The LD score intercept was obtained using snp_ldsc()in the LDpred2 package [16].

#### Polygenic score derivation

LDpred2-auto derives a PGS based on a prior for effect sizes of SNPs, GWAS summary statistics, and a LD matrix [17,18]. Using the bigsnpr package, we read dosages data from BGEN files using snp_readBEGN(). We used the 1,054,330 HapMap3 variants LD reference panel based on 362,320 European participants in UK Biobank provided by the authors [17,19]. We restricted SNPs to these HapMap3 variants and excluded any remaining variants that deviated from Hardy-Weinberg equilibrium (p<1e-7). We used snp_ldpred2_auto() to compute posterior effect sizes of selected SNPs, using 30 initial values for the proportion of causal variants from 1e-4 to 0.9 equally spaced on the log scale [19]. The initial heritability value for LDpred2-auto was obtained using constrained LD score regression. We specified 1,000 burn-in and 2,000 iterations for Markov Chain Monte Carlo method. Thirty PGS predictions were created in the independent sample (described in the next section) using the new effect sizes (Figure 1). After filtering the outlier predictions (>3 median absolute deviations from their median), we took the mean of the remaining predictions as the final PGS [19].

Fivefold cross-validation evaluated the PGS performance (77,770 participants randomly split into five equal sets of participants). GWAS and LDpred2-auto were performed in the training sets (i.e., combining 4 of the 5 split sets), and R^2^ (i.e., variance explained of device-measured sleep duration by the standardised PGS) was evaluated in the test set (Figure 1).

### Outcome ascertainment

UK Biobank provided the algorithmically defined outcomes, including all-cause dementia and three subtypes of dementia (category 460 [20]), combining data sources from self-reported verbal interview, hospital admission data, and death register data (https://biobank.ndph.ox.ac.uk/ukb/ukb/docs/alg_outcome_main.pdf). For each disease, the earliest date of the disease identified in any of the data sources was recorded. A previously published validation study of health record data found that the positive predictive values for Alzheimer’s disease were 68.2% in hospital admissions and 50% in mortality data [21].

We excluded participants with a prior history of any dementia at recruitment (determined by date of recruitment, data field 42018 “Date of all cause dementia”, and data field 42019 “Source of all cause dementia”). The outcome of the primary analysis was the first dementia diagnosis with mention of Alzheimer’s disease. We identified participants with the first diagnosis of any dementia during follow-up and recorded their diagnosis date (data field 42018 “Date of all-cause dementia report”). Among those, participants who had their first diagnosis of Alzheimer’s disease recorded on the same date (data field 42020 “Date of Alzheimer’s disease report”) were considered a case. Participants were right-censored on the first diagnosis date of dementia that did not mention Alzheimer’s disease (data field 42018 “Date of all cause dementia report”), date of death, date of lost to follow-up, or end of follow-up according to region at recruitment (England:30/9/2021;Scotland:31/7/2021;Wales:31/3/2016). As dementia usually presents in old age we conducted a sensitivity analysis restricted to participants aged >=60 years at recruitment, given our 12.5 median years of follow-up [22].

As a sensitivity analysis, we used a proxy for Alzheimer’s disease and related dementia (ADD) (hereafter called proxy-ADD) based on parental history of Alzheimer’s disease and related dementia as the outcome; this parental ADD proxy has been used in other GWASs to increase statistical power [23-25] and in MR [26], assuming that those with a family history have a higher number of risk variants than those without. We only considered parental history of ADD using self-reported data collected at recruitment (data fields 20107 and 20110). Individuals who were adopted as a child (data field 1767), who did not know whether their parents are alive or dead (data fields 1797 and 1835), or who reported “Do not know (group 1)” or “Prefer not to answer (group 1)” for data fields 20107 and 20110 were treated as “unknown” (ADD was included in group 1 in the question). After excluding participants with any unknown data on illnesses for both parents, a dementia-free participant who reported ADD in one or both natural parents was a proxy-ADD case (no weight adjustment for both affected parents).

Dementia-free participants who did not report any parental history of ADD at recruitment was a proxy-ADD control. The exclusion chart is shown in Figure 2.

**Figure 2.**
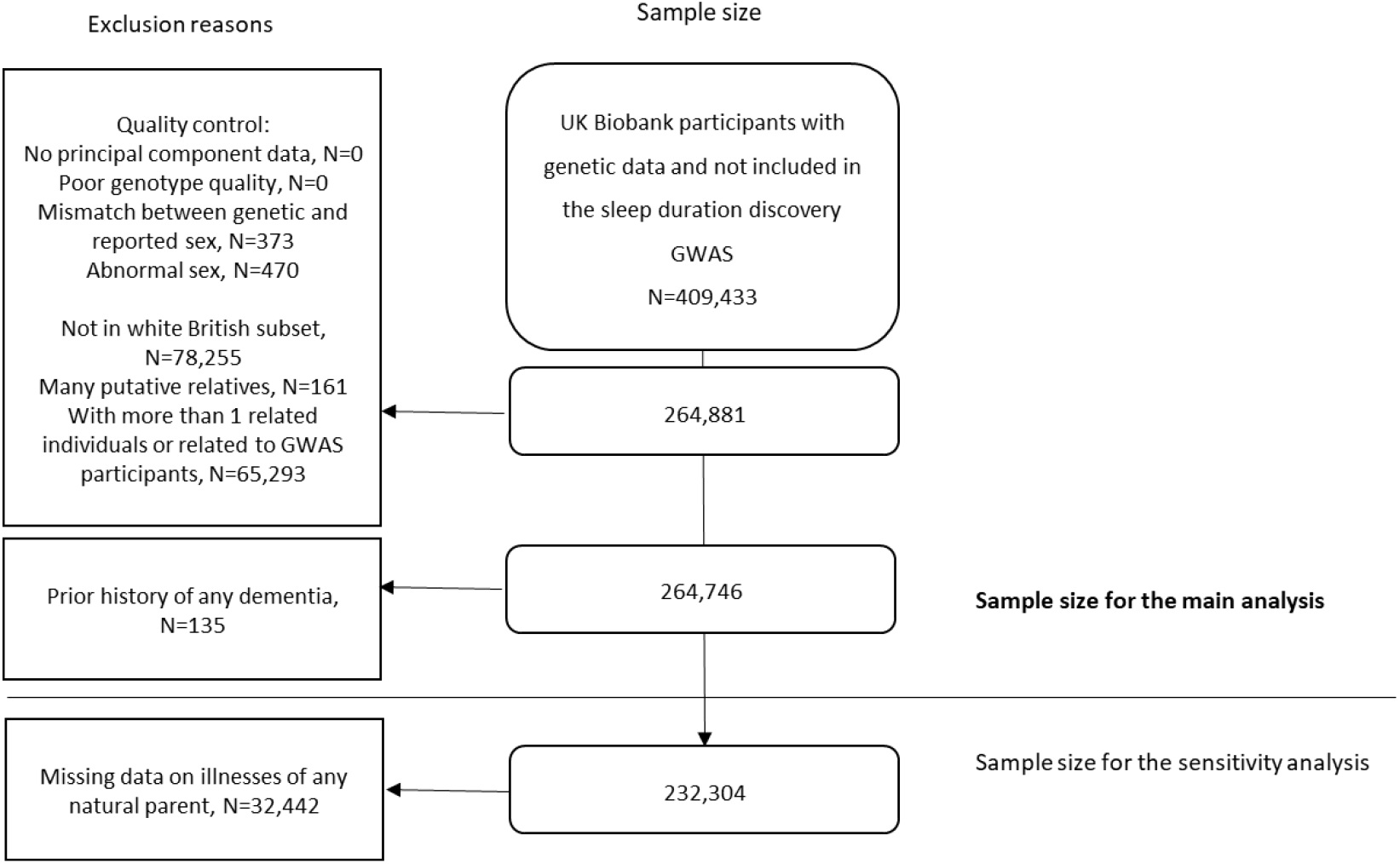
Flowchart of exclusion of participants for regression analyses with Alzheimer’s disease and with proxy for Alzheimer’s disease/dementia

## Statistical analyses

Among participants in UK Biobank who had imputed genotyping data, we applied the genetic quality control (QC) and restricted the sample to unrelated white British who also were not related to individuals included in the GWAS to derive our sleep PGS (Figure 2). Participants were categorised into fifths according to quintiles of the standardised sleep duration PGS using the new effect sizes obtained from LDpred2-auto in the independent 77,770 sample, with 40-60% (the middle fifth) as the reference group. Using GenoPred, we translated the PGS to the absolute scale in terms of predicted mean (95% CI) device-measured sleep duration [27], in order to assist interpretation of the findings.

In the main analysis, we assessed Alzheimer’s disease risk in relation to fifths of the PGS for sleep duration (with the middle fifth as the reference group) using Cox regression, with attained age as the underlying time variable. The model was adjusted for the first 10 principal components, genetic sex, and genotyping array. The significance level was set a 5%. We assessed evidence for an interaction between PGS sleep duration and *APOE* ε4 (with and without *APOE* ε4 determined by rs429358 and rs7412) in the main analysis with Alzheimer’s disease.

To investigate potential horizontal pleiotropy for our sleep duration PGS with Alzheimer’s disease, which means the association of PGS with Alzheimer’s disease risk acts through biological pathways other than, or in addition to, its influence on sleep duration, we examined the PGS for sleep duration with risk factors for dementia suggested in the 2020 report of the Lancet Commission (except for air pollution [an external factor] and brain traumatic injury [not available]). These included dichotomised variables related to education level, smoking, obesity, reported diagnosed/treated for hypertension, diabetes, alcohol consumption, physical activity, reported hearing problems, social isolation, and having depressive symptoms [1] (Categories listed in Table S.1). Complete-case logistic regression analyses were performed, adjusting for the same covariates as the primary analysis plus age at recruitment and squared age at recruitment. Bonferroni correction was applied to account for multiple testing (0.05/10=0.005). Where we observed evidence for an association between our sleep duration PGS and a potential risk factor for Alzheimer’s disease, we conducted additional analyses for sleep duration PGS and Alzheimer’s disease to assess whether our results may be sensitive to adjustment for these variables (with more refined categories in the additional analyses where appropriate). We also further assessed the PGS for sleep duration with self-reported sleep duration, as self-reported sleep duration was known to be genetically moderately associated with device-measured sleep duration in UK Biobank [28].

For the sensitivity analysis, we analysed only those participants with information on parental history of dementia, and estimated OR (95% CI) of proxy-ADD for the PGS for sleep duration using logistic regression, adjusting for the same covariates plus age and squared age at recruitment. To test linear trend, the value of the five PGS groups were set at 0.1, 0.3, 0.5, 0.7, and 0.9, respectively. Due to the late onset of dementia, we did a sensitivity analysis excluding participants whose parents were aged <60 years or whose parents died before reaching 60 years (data fields 2946 and 1845) or whose parents’ ages were unknown (responses of “Do not know” and “Prefer not to answer”) [24].

In the manuscript, floating absolute risks and 95% CIs were presented in figures [29], whereas conventional risk ratios and 95% CIs were presented in text and tables. Plotting was performed using Jasper in R [30].

## Results

### GWAS for sleep duration

Of the 77,770 participants included in the GWAS (Table 1), 34,564 were men (44%) and 43,206 (56%) were women with a mean age at wearable monitoring of 62.7 (SD, 7.8) years. The mean accelerometer-derived sleep duration was 8 hours 40 minutes (SD, 1.17h). GWAS results are presented in Manhattan and quartile-quartile plot (Figure S.1). While the lambda for genome inflation was 1.15, the LD score regression intercept (1.003) indicted that our results were likely due to a high polygenic trait and not biased by population stratification.

### Polygenic score performance

A total of 1,045,536 SNPs were included in the PGS for sleep duration. No SNPs were excluded based on the quality control of the summary statistics suggested by the authors [17]. SNP-heritability was 15.5% estimated by constrained LD score regression as required by LDpred2 [17]. The PGS explained 2.06% of the variance of total sleep duration in 5-fold cross validation. In the test set, the mean (SD) of the observed device-measured sleep duration within fifths of genetically predicted sleep duration by PGS were 8h25m (1.1 h), 8h35m (1.1h), 8h40m (1.2h), 8h45m (1.2h) and 8h54m (1.2h), respectively.

The regression analyses were conducted in an independent UK Biobank sample (N=264,746) that did not contribute the GWAS and PGS derivation. The PGS was constructed based on the numbers of risk alleles of the included SNPs and the final effect sizes obtained from LDpred2-auto. Using GenoPred with the mean and SD measured from the GWAS sample, the predicted mean (95% CI) sleep durations for the PGS fifths were 8h25m (6h9m-10h42m), 8h34m (6h18m-10h50m), 8h40m (6h23m-10h56m), 8h45m (6h29m-11h1m), and 8h54m (6h37m-11h10m) respectively (around 29 minutes device-measured sleep duration difference between the top and bottom fifths). Within the PGS fifths of the regression sample, the mean self-reported sleep durations were 7h5m, 7h8m, 7h10m, 7h12m, and 7h15m, respectively. In the adjusted linear regression (with adjustment for PC1-10, genotyping array, sex, age, squared age), the mean self-reported sleep duration in the top PGS fifth was 9.2 minutes longer than the bottom fifth.

Among the nine risk factors for dementia reported in the 2020 Lancet Commission report [1], a higher PGS indicating long sleep duration was inversely associated with having a college/university degree and with drinking alcohol daily/almost daily, and positively associated with hypertension and with reports of depressive symptoms (Figure S.2).

### Association analyses

The main analysis included 123,628 (47%) men and 141,118 (53%) women, and 1,451 Alzheimer’s disease cases were detected over a median 12.5 years of follow-up. Compared to the middle fifth, HRs(95%CI) for the 1^st^, 2^nd^, 4^th^ and 5^th^ fifths of the PGS sleep duration were 0.90(0.77-1.06), 0.97(0.83-1.14), 0.95(0.81-1.11), and 0.79(0.67-0.94), respectively (Figure 3). The sensitivity analysis restricting the sample to 121,610 participants aged 60 or above showed similar results: the HRs for the 1^st^, 2^nd^, 4^th^ and 5^th^ PGS fifths were 0.91(0.77-1.07), 0.96(0.81-1.13), 0.93(0.79-1.10), and 0.79(0.66-0.94), respectively. We did not find any evidence for effect modification of this association by *APOE* ε4 (p=0.48).

**Figure 3.**
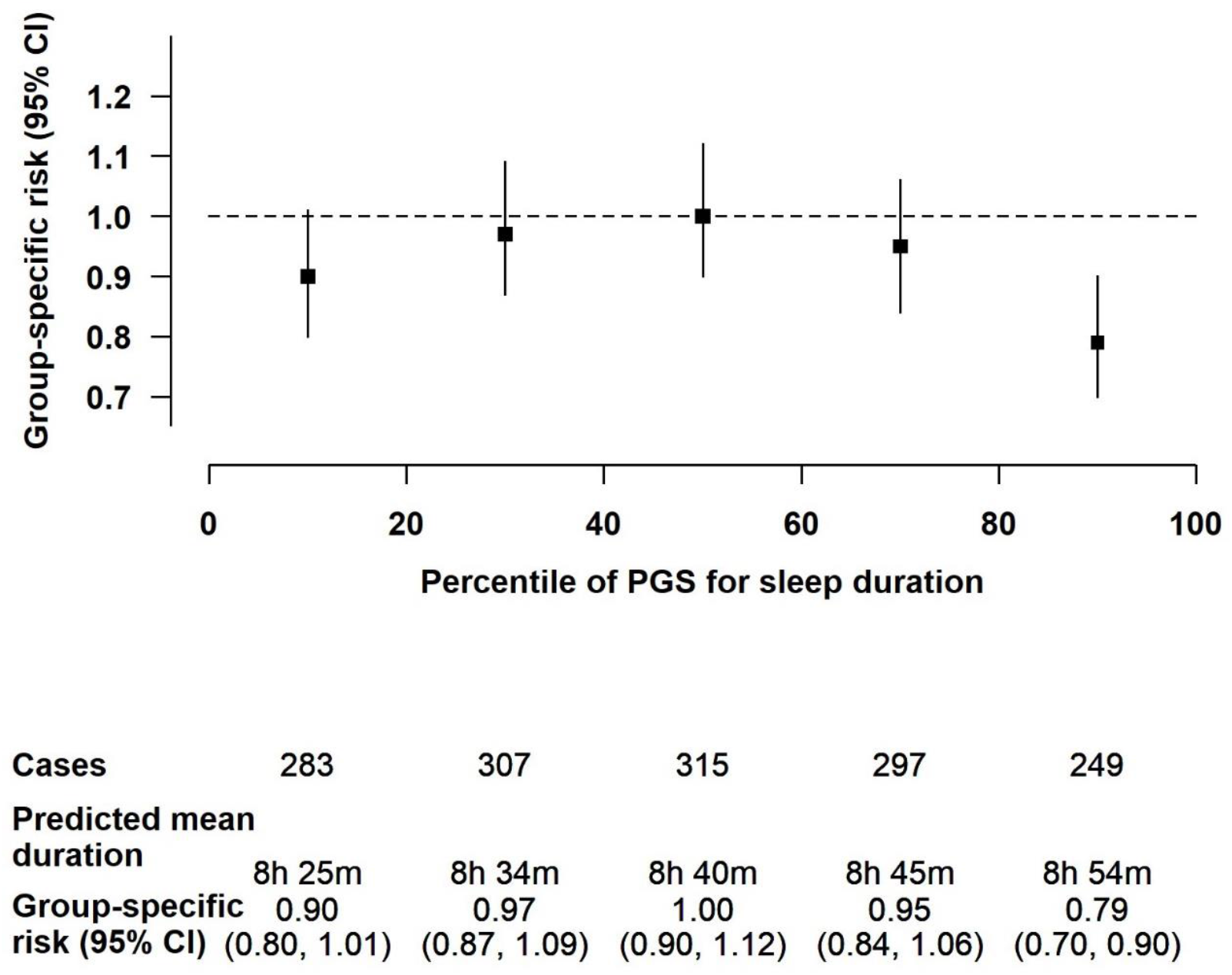
Association of the polygenic score for device-measured sleep duration with risk of Alzheimer’s disease in 264,746 UK Biobank participants The Cox model used attained age as the underlying time variable, and was adjusted for the first 10 principal components, genetic sex, and genotyping array

In view of the potential pleiotropic effects of the PGS sleep duration on educational attainment, hypertension, alcohol drinking frequency, and depressive symptoms, we made further adjustment for all these variables simultaneously in the main association analysis, but the results were not materially altered (Figure S.3).

In the sensitivity analysis using family history of Alzheimer’s disease/dementia, 28,520 out of 232,304 participants reported that one or both parents were affected by Alzheimer’s disease/dementia. Compared to individuals in the middle fifth of the PGS, those in the bottom two fifths had a marginally greater risk of proxy-ADD (Figure S.4). The ORs (95% CIs) for the 1^st^, 2^nd^, 4^th^ and 5^th^ fifths of PGS sleep duration were 1.04(1.00-1.09), 1.05(1.01-1.09), 1.01(0.97-1.05), and 1.00(0.96-1.04), respectively (p for trend=0.006). Analyses which restricted the age at baseline or age at death of participants’ parents yielded similar results: the ORs for the 1^st^, 2^nd^, 4^th^ and 5^th^ fifths of PGS sleep duration were 1.05(1.00-1.09), 1.05(1.01-1.10), 1.00(0.96-1.04) and 1.00(0.96-1.05), respectively.

## Discussion

We assessed sleep duration in relation to Alzheimer’s disease risk by leveraging a new genetic instrument for sleep duration, thus reducing the scope for reverse causation bias associated with observational measures of sleep duration. Using a recently published wearable-derived algorithm [11], our PGS sleep duration explained 6 times more variance (2.06% vs 0.33%) compared to that using significant SNPs in the discovery GWAS [4]. In the test set, people in the top fifth of the PGS had on average 29 minutes longer device-measured sleep duration than the bottom fifth. We observed that longer genetically predicted sleep duration was associated with a lower risk of Alzheimer’s disease risk.

Our findings provide a new line of evidence in favour of the potential role of sleep duration in Alzheimer’s disease risk. These findings contrast with multiple previously published MR studies that did not find evidence for an association between sleep duration and Alzheimer’s disease [6-8,31,32]. However, statistical power is notably limited in MR studies given the low variance explained by the genetic instruments used to predict sleep duration. A previously published study which used a PGS based method did not find an association between PGS for sleep duration with Alzheimer’s disease [33]. However, the PGS was based on self-reported sleep duration instead of the device-measured sleep duration we have used in the present study. Our sensitivity analysis suggested that shorter genetically predicted sleep duration, compared to the genetically predicted average sleep duration, was modestly associated with a higher risk of Alzheimer’s disease/dementia when using family history for ascertainment. This indicates compatibility with the hypothesis that sleep duration may be related to Alzheimer’s disease.

The associations of our PGS for device-measured sleep duration with frequent alcohol drinking, frequency of experiencing depressive symptoms, hypertension, and education attainment have not been reported elsewhere. The only exception was a MR analysis indicating self-reported sleep duration was inversely associated with hypertension [34]. While these factors did not appear to confound our observed association of the PGS with Alzheimer’s disease, it highlights the need for a more careful evaluation of whether these are confounding factors or mechanisms by which sleep duration is associated with Alzheimer’s disease. These factors may also be driven by genes that overlap with Alzheimer’s disease genes. For example, the genetic make-up of attained education and Alzheimer’s disease are strongly correlated [35].

### Strengths and limitations

This work represents another genetic approach to addressing reverse causation bias in the observational literature and the limited statistical power of GWAS significant genetic instruments for sleep duration. Other strengths of this study include a large sample size, the use of objective wearable measures, and the use of a PGS that explains much more trait variance (6x) compared to significant SNPs reported previously [4]. In future, replication of the association will be required in other cohorts to determine the utility of our device-measured sleep duration PRS in individuals of non-European genetic ancestry. One such example is the China Kadoorie Biobank [36] that has recently collected wrist-worn accelerometer and genomic data in 20,000 participants; and we would welcome similar data collections in other diverse biobanks.

## Conclusion

Our analyses indicate that longer genetically predicted average sleep duration is associated with lower Alzheimer’s disease risk. Our study also sheds light on the association of genetically predicted sleep and attained education, alcohol use, hypertension, and depression. Future research may benefit from disentangling how these factors explain the association between sleep duration and Alzheimer’s disease.

## Data Availability

Please refer to https://www.ukbiobank.ac.uk/ for details of UK Biobank.

## Supplementary Materials

**Table S.1.** Selected variables collected at recruitment for association analyses

| Variables | UK Biobank Data field | Categories used in the analysis | Notes (“Prefer not to answer”, “Do not know”, or missing data were regarded as unknown) |
| --- | --- | --- | --- |
| Education level | 6138 | Having a college/university degree, not having (ref.) |  |
| Hearing loss | 2247 | Yes, no (ref.) | Response of “I am completely deaf” are excluded from the analysis |
| Smoking | 20116 | Current or previous smoker, never smoker (ref.) |  |
| Hypertension | 6150, 20002, 6177, 6153 | Yes, no (ref.) | Reported a doctor diagnosis for hypertension (high blood pressure), reported having existing hypertension, or reported current use of blood pressure lowering medication were regarded as having hypertension; In data field 20002, ‘hypertension’ and ‘essential hypertension’ are considered as hypertension. |
| Frequent alcohol drinking | 1558 | Drinking daily or almost daily, less than daily/almost daily (ref.) |  |
| Obesity | 23104, 21001 | BMI≥30, <30 (ref.) kg/m<sup>2</sup> | The current measure is based on impedance measures (23104), if not calculated from weight and height measures (21001) |
| Depression symptoms | 2050 | At least several days (several days, more than half the days, nearly every day), not at all (ref.) |  |
| Social isolation | 709, 6160, 1031 | Score ≥2, score <2 (ref.) | A score was given for a response of “living alone” to this question: including yourself, how many people are living together in your household; A score was given for a response of frequency corresponding to |
|  |  |  | “about once a month” or less to this question: how often do you visit friends or family or have them visit you; A score was given for a response of “none of the above” to this question: which of the following (sports club or gym, pub or social club, religious group, adult education class, other group activity) do you attend once a week or more often [37]. |
| Physical activity | 894, 884, 914, 904 | Any moderate/vigorous activities of 2.5+ hours (or 150+ minutes) per week [38], <150 minutes (ref.) | Any missing in frequency/duration in moderate or vigorous activities were excluded from the analysis; Any reported duration of moderate/vigorous activities less than 10 minutes per day were recoded as 0 minutes. Any reported duration more than 180 minutes per day were recoded as 180 minutes. |
| Diabetes | 2443 | Yes, no (ref.) | Self-reported diabetes (we assumed the majority is type II diabetes) |
| Sleep duration | 1160 | No. of hours (possible values: 1-23 hours) |  |

**Figure S.1.**
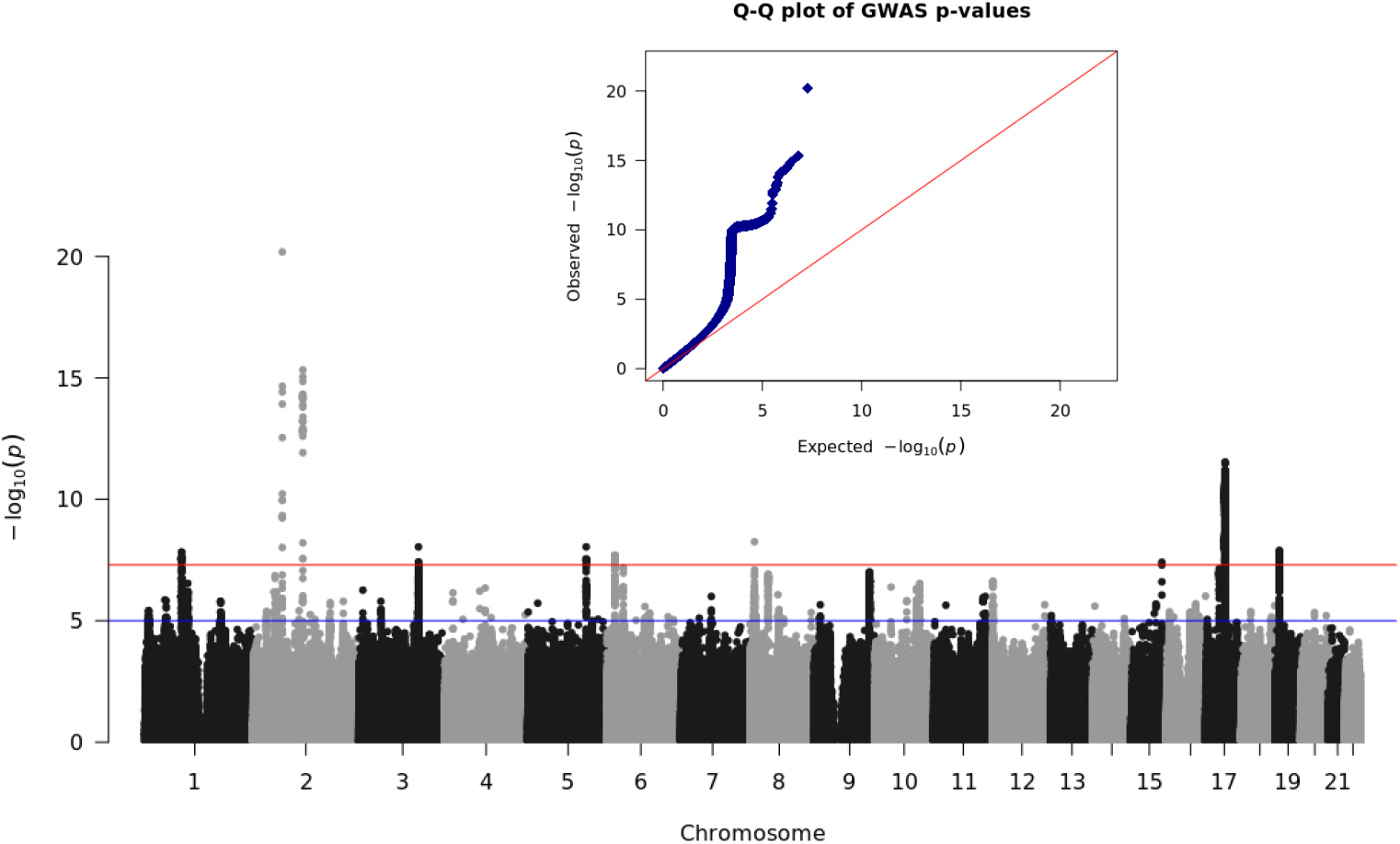
Manhattan plot of the genome-wide association study of device-measured sleep duration and quantile-quantile plot Lambda=1.15, LD intercept=1.003

**Figure S.2.**
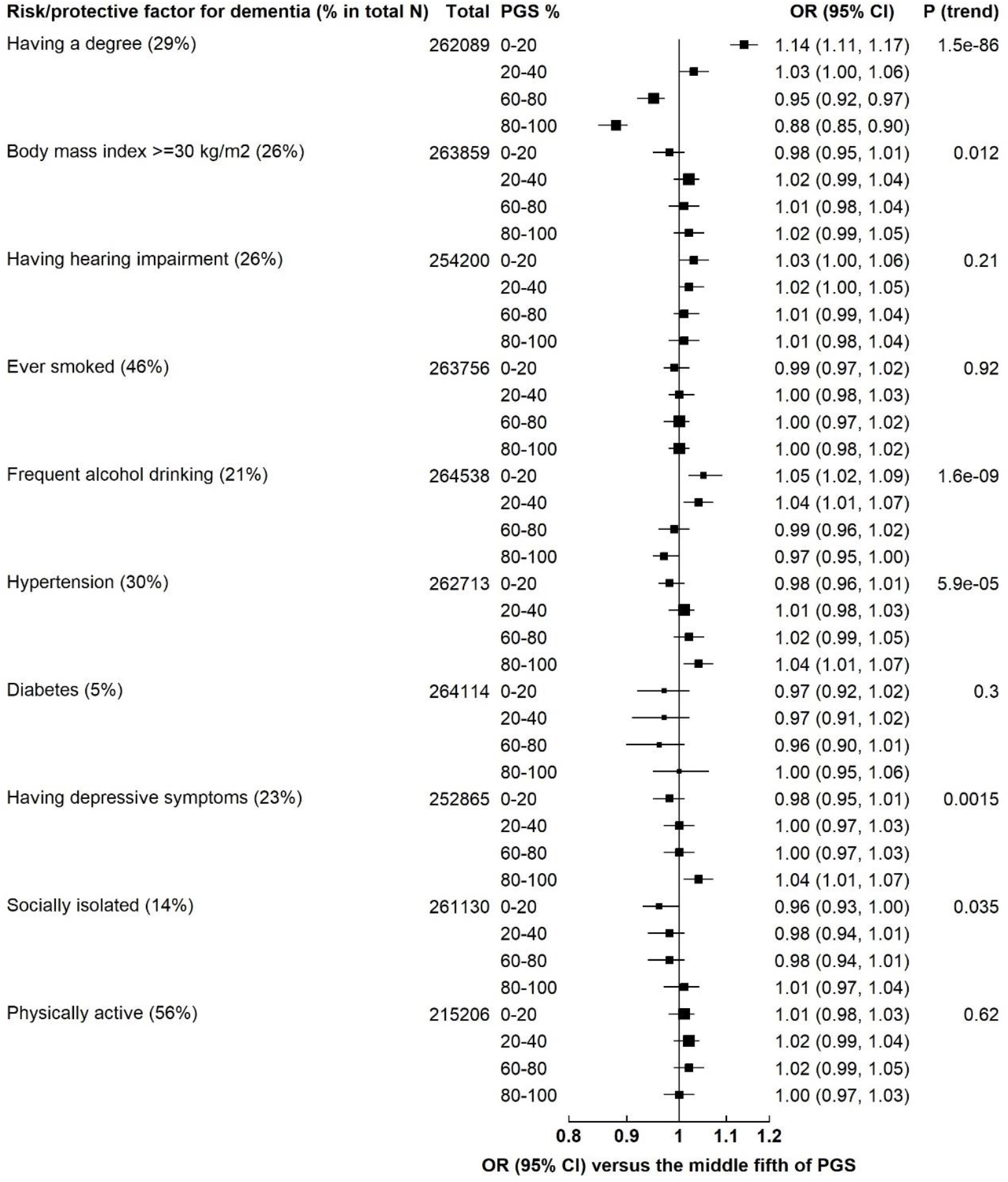
Odds ratios for proposed dementia risk/protective factors for non-baseline fifths versus the middle fifth of the polygenic score for sleep duration In this figure, statistical significance for trend tests was set at 0.005 after Bonferroni correction.

**Figure S.3.**
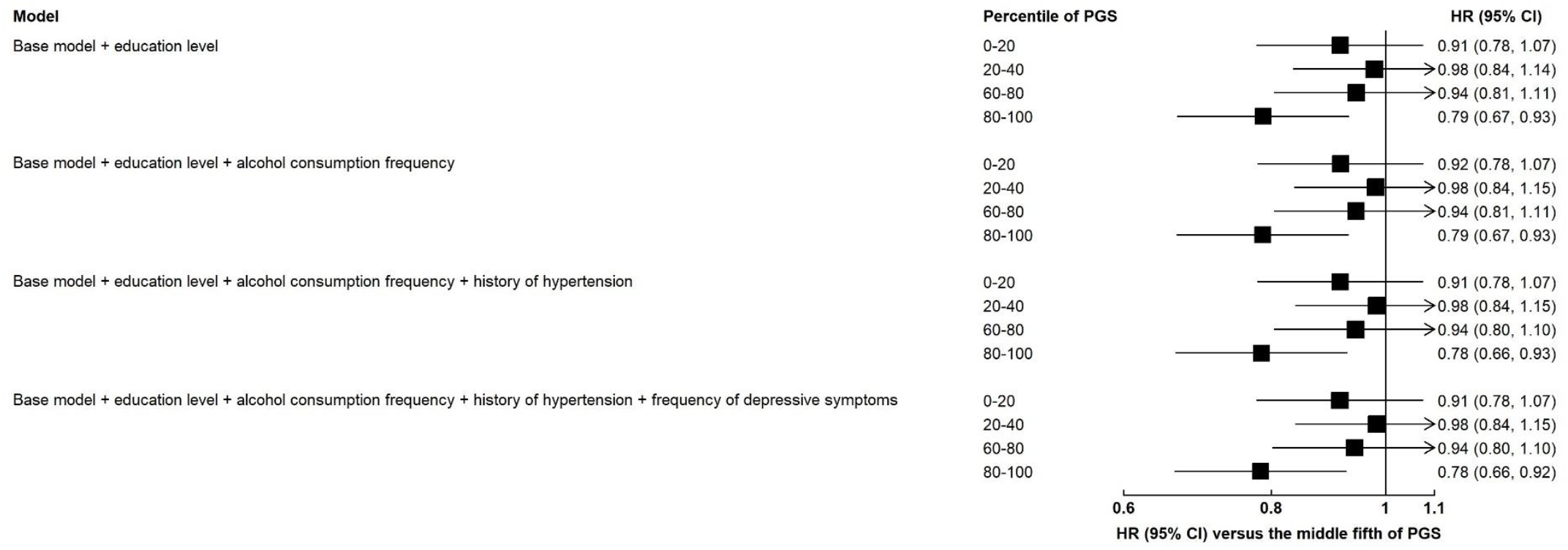
Additional adjustment for education, alcohol drinking frequency, history of hypertension, and frequency of depressive symptoms to assess the sensitivity of the multivariable-adjusted association between the polygenic score for sleep duration and Alzheimer’s disease risk to pleiotropic factors Note: The reference group is the 40-60% of the polygenic score for sleep duration. The base model used attained age as the underlying time variable, and was adjusted for the first 10 principal components, genetic sex, and genotyping array. Categories for the further adjustment variables: education level (higher education (having a college or university degree as the highest qualification), further education (having A levels/AS levels or equivalent, NVQ or HND or HNC or equivalent, or other professional qualifications e.g., nursing, teaching as the highest qualification), school leaver (CSEs or equivalent or O levels/GCSEs or equivalent as the highest qualification, or none of the above), unknown), alcohol consumption frequency (never, <3 times per week, 3+ times per week, unknown), history of hypertension (yes, no, unknown), and frequency of depressive symptoms (not at all, several days, more than half the days or nearly every day, unknown). PGS: Polygenic score.

**Figure S.4.**
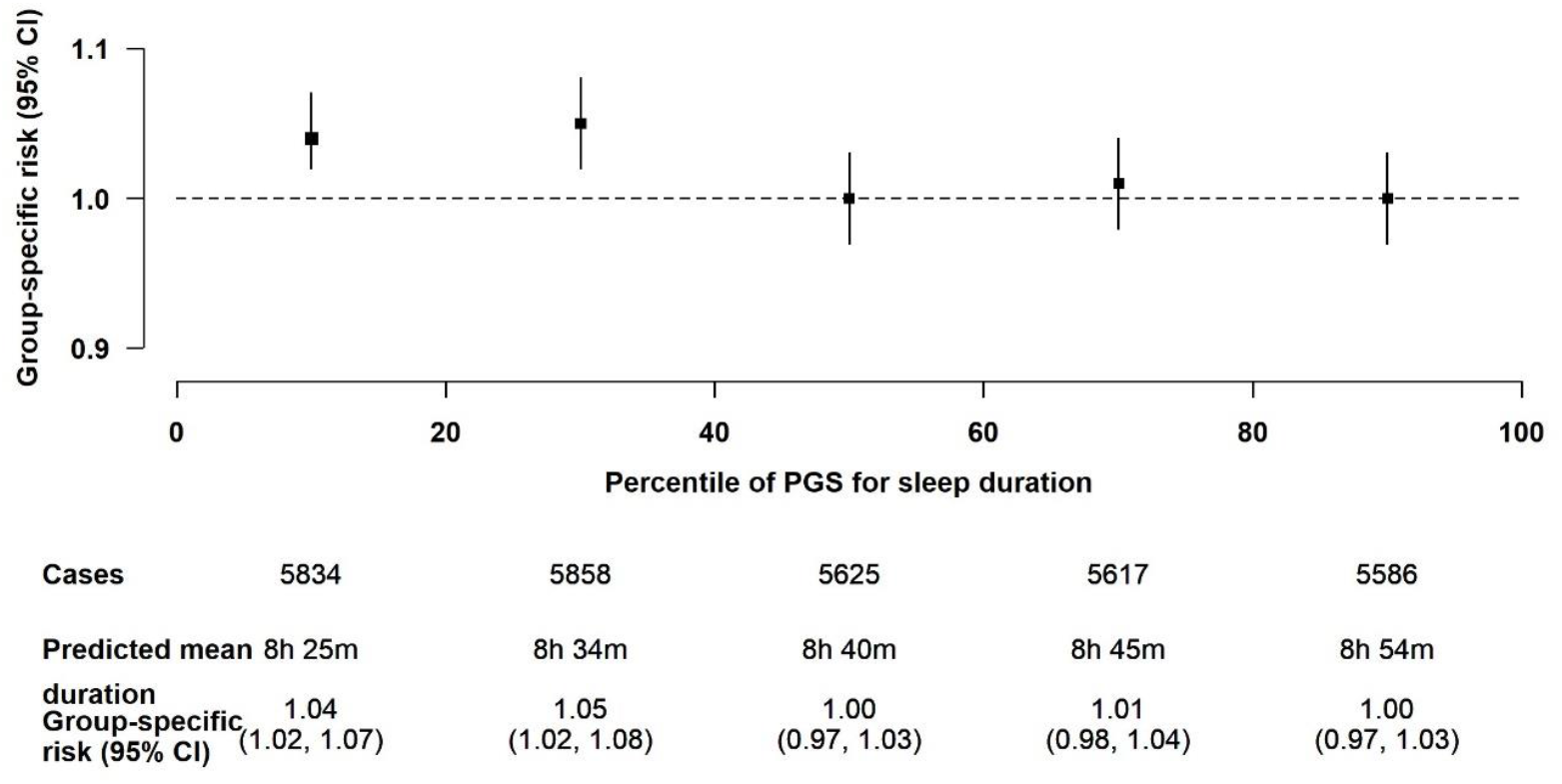
Association of the polygenic score for sleep duration with odds of having a family history of Alzheimer’s disease/dementia in 232,304 participants in UK Biobank who had data on history of illnesses of their parents The model was adjusted for the first 10 principal components, genetic sex, genotyping array, age at recruitment, and squared age at recruitment.

## Acknowledgment and Funding statement

The UK Biobank application associated with the current study is 59070. ATYW is supported by a Nuffield Department of Population Health (NDPH) Early Career Research Fellowship. AD is supported by the Wellcome Trust [223100/Z/21/Z], Novo Nordisk, Swiss Re, the National Institute for Health Research (NIHR) Oxford Biomedical Research Centre (BRC), the British Heart Foundation Centre of Research Excellence (grant number RE/18/3/34214), the Alan Turing Institute and the British Heart Foundation (grant number SP/18/4/33803), and Health Data Research UK, an initiative funded by UK Research and Innovation, Department of Health and Social Care (England) and the devolved administrations, and leading medical research charities. KSB is funded by The Cancer Research UK Programme grant (C8221/A29017). SF and GKR are PIs of the Million Women Study which is funded by Cancer Research UK (A29186). MVH was supported by a British Heart Foundation Intermediate Clinical Research Fellowship (FS/18/23/33512). RCT is funded by Cancer Research UK (C8221/A29017). Computation used the Oxford Biomedical Research Computing (BMRC) facility, a joint development between the Wellcome Centre for Human Genetics and the Big Data Institute supported by Health Data Research UK and the NIHR Oxford Biomedical Research Centre. The views expressed are those of the author(s) and not necessarily those of the NHS, the NIHR or the Department of Health.

